# A genome-wide association study of Chinese and English language abilities in Hong Kong Chinese children

**DOI:** 10.1101/2022.08.01.22278296

**Authors:** Yu-Ping Lin, Yujia Shi, Ruoyu Zhang, Xiao Xue, Shitao Rao, Kevin Fai-Hong Lui, Dora Jue Pan, Urs Maurer, Richard Kwong-Wai Choy, Silvia Paracchini, Catherine McBride, Hon-Cheong So

## Abstract

**Background:** Reading and language skills are important and known to be heritable, and dyslexia and developmental language disorder are commonly recognized learning difficulties worldwide. However, the genetic basis underlying these skills remains poorly understood. In particular, most previous genetic studies were performed on Westerners. To our knowledge, few or no previous genome-wide association studies (GWAS) have been conducted on literacy skills in Chinese as a native language or English as a second language (ESL) in a Chinese population.

**Methods:** We conducted GWAS and related bioinformatics analyses on 34 reading/language-related phenotypes in Hong Kong Chinese bilingual children (including both twins and singletons; *N*=1046). We performed association tests at the single-variant, gene, pathway levels, and transcriptome-wide association studies (TWAS) to explore how imputed expression changes might affect the phenotypes. In addition, we tested genetic overlap of these cognitive traits with other neuropsychiatric disorders, as well as cognitive performance (CP) and educational attainment (EA) using polygenic risk score (PRS) analysis.

**Results:** Totally 9 independent loci (LD-clumped at r^2^=0.01) reached genome-wide significance (*p*<5e-08) (filtered by imputation quality metric Rsq>0.3 and having at least 2 correlated SNPs(r^2^>0.5) with *p*<1e-3). The loci were associated with a range of language/literacy traits such as Chinese vocabulary, character and word reading, dictation and rapid digit naming, as well as English lexical decision. Several SNPs from these loci mapped to genes that were reported to be associated with intelligence, EA other neuropsychiatric phenotypes, such as *MANEA, TNR, PLXNC1* and *SHTN1*. We also revealed significantly enriched genes and pathways based on SNP-based analysis. In PRS analysis, EA and CP showed significant polygenic overlap with a variety of language traits, especially English literacy skills. ADHD PRS showed a significant association with English vocabulary score.

**Conclusions:** This study revealed numerous genetic loci that may be associated with Chinese and English abilities in a group of Chinese bilingual children. Further studies are warranted to replicate the findings and elucidate the mechanisms involved.

## Introduction

Literacy and language skills are important for academic development in children. Learning difficulties (e.g., dyslexia) are common and may affect one’s school performance, leading to poorer work attainment and socioeconomic status, as well as decreased general well-being ^1^. Multiple cognitive and language skills often serve as a strong foundation for literacy and language development; these include working memory, rapid naming, and vocabulary knowledge ^2^. Different factors of environmental and genetic origins may also affect children’s literacy-related and language-related skills across languages. Family, twin, and adoption studies have provided strong evidence that these complex cognitive and language traits and academic performance in young children are heritable ^3 4 5 6 7^ and also highly polygenic ^8 9^. However, the exact genes/variants involved in these traits are still not well understood, probably due to the complexity of the phenotypes and difficulty in gathering sufficient samples.

In recent years, several GWAS studies have been conducted on reading and language abilities in European populations. Several studies have focused on developmental dyslexia (DD) or high/low reading ability as a binary outcome, adopting a case-control study method ^9 10 11 12 13 14^. Such study design may enable a larger sample size to be collected, but also has its shortcomings. Language and literacy skills cover a broad range of phenotypes, and dyslexia is also a highly heterogenous condition. The focus on a single binary outcome may limit our understanding into the biological mechanisms underlying different domains of language abilities. Other studies have investigated reading and language abilities as continuous traits ^8 15 16 17 18 19 20^.

However, given the relatively high heritability of literacy and language skills ^21 22^, the genetic variants discovered thus far are still far from explaining the full genetic basis of these complex traits. In addition, most previous GWAS were conducted in European populations. However, the genetic architecture of language phenotypes may be different across ancestries, and some of the variants may be more readily discovered in other populations due to differences in allele frequency or LD (linkage disequilibrium) structure.

In addition, to our knowledge, no previous GWAS have been published on Chinese children’s literacy and language skills in native Chinese and English as a second language (ESL). We note that in one recent GWAS on dyslexia ^9^, several associated loci were also replicated in the Chinese Reading Study of reading accuracy and fluency; yet the primary GWAS was conducted predominantly on population of European ancestry. Given possible differences in mechanisms underlying Chinese and English literacy and language skills, it is essential to study the genetic variants underlying Chinese literacy phenotypes. While some previous studies have investigated the genetics of cognitive and language phenotypes, most have only focused on a limited number or domain of phenotypes (e.g., rapid naming, word reading).

In view of the above limitations, here we conducted GWAS and related bioinformatics analyses on a comprehensive panel of 34 literacy/language-related phenotypes in a Hong Kong Chinese population. The wide coverage enables a systematic and unbiased analysis of a variety of literacy and language-related phenotypes. Since this is among the first study of Chinese- and ESL-related phenotypes in a Chinese population, and the genetic bases of such phenotypes are still largely unknown, it is our objective to explore a wider range of traits to maximize the chance of discovery, and to provide a starting point and also important reference for future studies.

Briefly, in this work, we investigated how genetics is associated with individual differences in Chinese and English reading and writing. We performed association tests at the single-variant, gene, and pathway levels, and employed transcriptome-wide association studies (TWAS) to explore how genotype-imputed expression changes affect the phenotypes. In addition, we tested potential associations between these complex cognitive traits with other neuropsychiatric disorders, as well as cognitive performance and educational attainment by polygenic risk score (PRS) analysis. To the best of our knowledge, this is the first GWAS conducted on a comprehensive range of Chinese-language and ESL-related phenotypes in a Chinese population.

## Subject and methods

### 1. Participants

The participants in our study were Hong Kong Chinese-English bilingual twins and singletons, with Chinese (Cantonese) as their native language. Children aged between 3 to 11 were recruited through kindergarten and primary schools in Hong Kong. A total of 1048 children were recruited for this genetic study, including 274 MZ subjects (137 pairs), 350 DZ subjects (175 pairs) and 424 singletons. Zygosity determination on twin pairs was based on the genotyped small tandem repeat (STR) markers using Quantitative Fluorescence Polymerase Chain Reaction (QF-PCR)^23^. Singleton children were selected from the same schools as those twin pairs. Parental written consent for all the participants was obtained before testing. Children completed a series of cognitive and literacy-related tasks in Chinese and English either in a laboratory setting, their school, or their home by trained research assistants. For details of the tasks and phenotypes, please refer to the supplementary text. Briefly, a total of 34 phenotypes were included (Table 1), covering a wide range of literacy- and language-related skills.

**Table 1.** Overview of phenotypes included in the study.

| **Variable** | **Variable Label** |
| --- | --- |
| BDS_Total | Backward Digit Span |
| CCR_Total | Chinese Character Reading |
| CDC_Total | Chinese Delayed Copying |
| CDICT_Total | Chinese Dictation |
| CDRAN_Mean | Chinese Digit Rapid Naming |
| CLD_Total | Chinese Lexical Decision |
| COM_Score | Chinese 1 Min Word Reading Adjusted Total Score |
| COM_Norm | Chinese 1 Min Word Reading Scaled Score |
| CVA_Total | Chinese Vocabulary - Receptive Vocabulary (10 items) |
| CVB_Total | Chinese Vocabulary - Expressive Vocabulary (12 items) |
| CVD_Total | Chinese Vocabulary - Vocabulary Definition (26 items) |
| CVK_Total | Chinese Vocabulary Knowledge (48 items; sum of CVA, CVB and CVK) |
| CWR_Total | Chinese Word Reading Raw Score |
| CWR_Norm | Chinese Word Reading Scaled Score |
| DS_Total | Chinese Discourse Skills |
| EDC_Total | English Delayed Copying |
| EDICT_Total | English Dictation |
| EDRAN_Mean | English Digit Rapid Naming |
| EIS_Total | English Invented Spelling |
| ELD_Total | English Lexical Decision |
| ELRAN_Mean | English Letter Rapid Naming |
| EMA_Total | English Morphological Awareness - Written Test |
| EVA_Total | English Vocabulary - Receptive Vocabulary (15 items) |
| EVB_Total | English Vocabulary - Expressive Vocabulary (15 items) |
| EVD_Total | English Vocabulary - Vocabulary Definition (15 items) |
| EVK_Total | English Vocabulary Knowledge (45 items; sum of EVA, EVB and EVK) |
| EWR_Total | English Word Reading Total Score |
| MS_Total | Morphosyntax |
| PairC_Total | Pair Cancellation |
| PureC_Total | Pure Copying of Unfamiliar Scripts |
| RC_MC | Reading Comprehension - Multiple Choice |
| RC_OE | Reading Comprehension - Open End |
| RC_Total | Reading Comprehension - Total |
| WO_Total | Chinese Word Order |

All tasks were finished in a given order that had been predetermined. Please refer to the supplementary text for a detailed description of each studied phenotype. A correlation matrix of all phenotypes is presented in Figure S1.

### 2. Genotype quality control (QC) and imputation

Three groups of subjects, including monozygotic (MZ) twins, dizygotic twins (DZ), and singletons, were genotyped. Based on previous studies ^24^, reducing the MZ pairs to singletons leads to a loss of statistical power. It has also been shown that including both MZ twins in the genetic analysis does not lead to an inflation of type I error (when relatedness is accounted for) but can improve power ^24^. We therefore followed ref ^24^ and included both MZ twins in our GWAS. Monozygosity was confirmed by QF-PCR as described above, and only one member of each MZ pair was genotyped. The other MZ twin was assumed to share identical genotypes.

Quality control (QC) was performed by PLINK-1.9 on each dataset separately before merging. We removed those SNPs which deviated from Hardy–Weinberg equilibrium (HWE, P < 1E-5), with Minor Allele Frequency (MAF) < 1%, missingness per individual (MIND) > 10%, and missingness per marker (GENO) > 10%. After QC, 911178 SNPs and 1046 individuals were kept for further analysis, including 274 MZ subjects (59 male pairs, 78 female pairs), 349 DZ subjects (39 male pairs, 37 female pairs, 1 member of a female pair and 98 opposite-sex pairs), as well as 423 singletons (218 males, 205 females).

Following QC, variant-level imputation was performed by the Michigan Imputation Server based on “Mininac” ^25^. The imputation was based on the reference panel 1000 Genomes (1000G) Phase 3 v5, as previous studies reported satisfactory performance of imputation in Chinese based on the 1000G panel ^26^. The imputed data were converted into a binary dosage file by the program “DosageConverter” (https://genome.sph.umich.edu/wiki/DosageConvertor). Imputed variants with INFO score (R-squared) > 0.3 (12,475,316 SNPs) were retained.

### 3. Genome-wide association study

A genome-wide association study (GWAS) of all phenotypes was conducted through a univariate linear mixed model in GEMMA (http://github.com/genetic-statistics/GEMMA). We included age and sex as fixed-effects covariates. The genetic relationship matrix (GRM) was included as a random effect to account for relatedness between subjects. This approach also controls for population stratification. We tested for the association of allelic dosages with phenotypes as described above. We considered *p*<5e-8 as the genome-wide significance threshold. Although multiple phenotypes were studied, our primary objective was to explore and prioritize genetic variants for further studies, and a further Bonferroni correction to penalize the number of phenotypes tested may be too conservative for this purpose. Instead, we employed the false discovery rate (FDR) approach to control for multiple testing. FDR controls the expected *proportion* of false positives among the results declared to be significant. This approach has been argued to be a more reasonable methodology as it ‘adaptively’ considers the data instead of imposing a direct penalty for the number of hypotheses tested, and the FDR approach has also been widely used in genomic studies ^27^.

To identify independent significant risk loci, we employed PLINK-1.9 to perform LD-clumping with r^2^=0.01 and distance = 1000kb, using 1000 Genomes East Asian sample as reference. SNP-to-Gene mapping was done using Bioconductor’s software package ‘biomaRt’ (version 2.48.2) on R-4.0.3.

Histograms of all phenotypes are shown in Figure S2; some of the phenotypes were normally distributed though some were not. Nevertheless, in large sample sizes with few covariates, violation of the normality assumption often does not affect the validity of results ^28^. There is no clear consensus on whether transformations (such as the rank-based inverse normal transformation, RINT) should be performed on (non-normal) phenotypes in GWAS. For example, Beasley et al. ^29^ reported that RINT does not necessarily control type I error and may lead to reduced statistical power, while another study ^30^ showed improved performance of the RINT approach. Intuitively, the untransformed approach keeps the original value of the phenotype and does not lead to loss of information, and is more interpretable. Here we performed analysis on both RINT-transformed ^30^ and non-transformed phenotypes for all traits under study. As described below, on inspection of the QQ-plots, most traits have very similar distributions of p-values, except for four phenotypes. We primarily present our results of the non-transformed phenotypes except for the latter four which were RINT-transformed.

### 4. Gene-based test and pathway enrichment analysis

#### 4.1 Gene-based analysis with MAGMA

Gene-based analysis has been considered more powerful than SNP-based analysis performed in GWAS ^31^. We utilized MAGMA (Multi-marker Analysis of GenoMic Annotation) v1.06 to conduct gene-based association tests with GWAS summary statistics of our phenotypes ^13^. Briefly, MAGMA considers the aggregate effects of all variants in each gene to produce a gene-based test statistic. We employed the FDR procedure ^32^ to control for multiple testing. In our gene-based study and the following analyses, results with FDR < 0.05 are regarded as significant, while those with 0.05 < FDR < 0.2 are considered to be suggestive associations.

#### 4.2 Pathway analysis with GAUSS

We subsequently performed pathway enrichment tests with a powerful subset-based gene-set analysis method called GAUSS (Gene-set analysis Association Using Spare Signal) ^33^, based on gene-based association results obtained by MAGMA. We utilized two collections of gene-sets derived from the Molecular Signature Database (MsigDB v6.2) ^34^. The first is a collection of curated pathways (C2) which include canonical pathways such as KEGG, BioCarta, REACTOME, as well as chemical and genetic perturbations; the other is gene-ontology (GO) gene-sets (C5), which include biological processes, molecular processes, and cellular processes. Please refer to https://www.gsea-msigdb.org/gsea/msigdb/collections.jsp for details. If a significant association with a pathway is found, GAUSS also identifies the core subset (CS) of genes within the pathway that is driving the association. FDR was used to control for multiple testing.

#### 4.3 Transcriptome-wide association studies with S-Predixcan & S-Multixcan

We have also employed other approaches to compute gene-based association results. MAGMA is a widely used approach, but it does not consider the functional impact of SNPs (e.g., impact on expression). S-PrediXcan is another gene-based analysis approach which *imputes* gene expression changes in relevant tissues due to genetic variations, using reference eQTL datasets such as GTEx. This approach is also known as the transcriptome-wide association study (TWAS) ^35^. Here we considered 13 brain regions, including the amygdala, anterior cingulate cortex (BA24), caudate basal ganglia, cerebellar hemisphere, cerebellum, cortex, frontal cortex (BA9), hippocampus, hypothalamus, nucleus accumbens (basal ganglia), putamen (basal ganglia), spinal cord (cervical c-1) and substantia nigra. To increase statistical power to identify candidate genes, we integrated the joint effects of expression changes across multiple tissues in a secondary analysis by ‘S-MultiXcan’ ^36^. S-MultiXcan combines evidence across tissues using multiple regression (fitting predicted expression as independent variables), which also takes into account the correlation structure.

### 5. Polygenic risk score analysis

To evaluate the genetic overlap of the studied phenotypes with other neuropsychiatric traits, we also performed a PRS analysis. A PRS analysis for individuals aggregates the joint effect of multiple genetic variants, weighted by the effect size from GWAS summary statistics data. PRS were generated by PLINK 1.9 across 11 P-value thresholds (pthres)={1e-06, 1e-05, 1e-04, 0.001, 0.01, 0.1, 0.2, 0.3, 0.4, 0.5,0.05}(multiple testing corrected by FDR) ^37^, LD-clumped at r^2^=0.1 within a distance of 1000 kb.

The neuropsychiatric traits for constructing PRS included educational attainment (EA) (N= 1,131,881) ^38^, cognitive performance (CP; N=257,841; derived from scores of verbal-numerical reasoning from the UK Biobank and neuropsychological test results from the COGENT Consortium, details described in ^38^), autism spectrum disorders (ASD; N=10610)^39^, attention deficit hyperactivity disorder (ADHD), (ADHD; N=55374) ^40^, schizophrenia (SCZ; N=105318)^41^, bipolar disorder (BP; N= 416053)^42^ and major depressive disorder (MDD; N=18759) ^43^. GWAS summary statistics were downloaded from the Social Science Genetic Association Consortium (SSGAC) (https://www.thessgac.org/), Psychiatric Genomics Consortium (PGC) (https://www.med.unc.edu/pgc) and The Integrative Psychiatric Research project (iPSYCH) (https://ipsych.au.dk/downloads/).

We employed linear mixed models in GEMMA to test for associations between PRS and phenotypes. The model was adjusted for age and sex as fixed effects. GRM was fit as a random effect, accounting for both relatedness and population stratification ^44^.

## Results

### Single-variant associations

Quantile-quantile plots (QQ-plots) with lambda (λ) were constructed for each trait with and without RINT transformation. We found that the QQ-plots were very similar for most phenotypes with and without the transformation, except for four [Backward digit span (BDS_Total), Chinese Vocabulary - Receptive Vocabulary (CVA_Total), Chinese digit rapid naming (CDRAN_Mean) and English digit rapid naming (EDRAN_Mean)] (see Figure S3 and Figure S4). For these 4 traits, subsequent analyses were based on the RINT-transformed values, and all four traits showed no evidence of inflated false positives after the transformation based on the updated QQ-plots. Manhattan plot for all traits are shown in Figure S5.

In SNP-based analysis, a total of 9 independent loci (LD-clumped at r^2^ threshold=0.01) reached genome-wide (GW) significance (*p* < 5e-08) with imputation quality score (Rsq)>0.3 and at least 2 correlated SNPs (r^2^>0.5) with *p*<1e-3 (Table 2). The check for correlated significant SNPs was performed to further reduce the risk of false positives, and was done using the default settings of LD-clumping in PLINK. The loci were associated with a variety of language/literacy traits such as Chinese vocabulary, character and word reading, dictation and digit rapid naming, as well as English lexicon decision. Note that 3 loci were each associated with two (correlated) phenotypes, including rs77868538 which was associated with both CVK_Total and CVD_total (correlation (r)=0.97), rs182977703 which was associated with both COM_Norm and CDICT_Total (r=0.37), and rs4865143 which was associated with CWR_total and CVB_total (r=0.63).

**Table 2.** Results of the SNP-based association analysis.

| Phenotype | CHR | BP | SNP | A1 | A2 | P | MAF | Rsq | Genotyped | Closest gene | S0001 | FDR-adjust *P* |
| --- | --- | --- | --- | --- | --- | --- | --- | --- | --- | --- | --- | --- |
| ELD_Total | 6 | 95643248 | rs6905617 | C | A | 3.29E-09 | 0.352 | 0.52 | Imputed | *MANEA-AS1(-364.7kb)* | 43 | 3.19E-03 |
| CVK_Total | 1 | 175436068 | rs77868538 | T | C | 3.33E-09 | 0.028 | 0.99 | Imputed | *TNR(0)* | 4 | 1.32E-02 |
| CVD_Total | 1 | 175436068 | rs77868538 | T | C | 7.44E-09 | 0.028 | 0.99 | Imputed | *TNR(0)* | 4 | 2.87E-02 |
| ELD_Total | 1 | 238306117 | rs370871011 | T | G | 9.34E-09 | 0.011 | 0.64 | Imputed | *LOC100130331(+214.5kb)* | 2 | 6.26E-03 |
| CCR_Total | 9 | 115640979 | rs56024259 | G | A | 1.53E-08 | 0.124 | 0.98 | Imputed | *SLC46A2(-0.22kb)* | 7 | 3.97E-02 |
| ELD_Total | 5 | 32421181 | rs925214 | T | G | 1.64E-08 | 0.030 | 0.86 | Imputed | *ZFR(0)* | 37 | 8.44E-03 |
| CDRAN_Mean | 12 | 94529190 | rs3847795 | A | C | 1.73E-08 | 0.173 | 0.94 | Imputed | *PLXNC1(-13.31kb)* | 4 | 7.60E-02 |
| ELD_Total | 1 | 202430424 | rs12049280 | G | T | 3.17E-08 | 0.021 | 0.66 | Imputed | *PPP1R12B(0)* | 5 | 1.52E-02 |
| CWR_Total | 4 | 57573275 | rs4865143 | T | C | 3.61E-08 | 0.071 | 0.80 | Imputed | *HOPX(+25.4kb)* | 15 | 1.34E-01 |
| COM_Norm | 10 | 118659171 | rs182977703 | G | T | 4.39E-08 | 0.016 | 0.60 | Imputed | *SHTN1 (0)* | 4 | 1.49E-01 |
| CDICT_Total | 10 | 118659171 | rs182977703 | G | T | 4.44E-08 | 0.016 | 0.60 | Imputed | *SHTN1 (0)* | 2 | 1.52E-01 |
| CVB_Total | 4 | 57573275 | rs4865143 | T | C | 4.97E-08 | 0.071 | 0.80 | Imputed | *HOPX(+25.4kb)* | 27 | 9.92E-02 |

We also provide the full list of all GW significant SNPs (with Rsq>0.3) in Table S1 (Table S1.1: all GW-significant hits; Table S1.2: LD-clumped significant hits; Table S1.3: all GWAS results with FDR<0.1). In addition, we also searched the top-listed genes in GWAS catalog for associations with other phenotypes (especially neuropsychiatric traits) in previous studies. Please refer to Table S10 for details.

We detected the largest number of genome-wide significant SNPs with English Lexical Decision (ELD) (see Table S1.2). The most significant association was observed for rs6905617 (C/A, MAF = 0.35, *p* = 3.29E-09) with ELD; the SNP is located close to *MANEA* (-382.1 kb) and *MANEA-AS1* (-364.7kb). As for Chinese-related traits, the strongest single-variant association with CVK (Chinese Vocabulary – Knowledge) was detected at rs77868538 (T/C, MAF = 0.02548, *p* = 3.33E-09), an imputed variant located within *TNR* (Tenascin R). In addition to CVK, we also discovered one significant locus for CCR, CWR, CDICT, CDRAN, COM, CVB and CVD respectively (Table 2).

We also calculated the lambda-GC (genomic inflation factor) for each untransformed trait and there was no evidence of inflation (Table S7; largest lambda-GC=1.0255, 29/34 traits showed lambda-GC<1.02).

### Association analyses between genetically predicted expression and phenotypes

We evaluated the association between genetically regulated expression (GRex) and phenotypes across multiple brain regions by S-Predixcan. We used pre-computed weights provided by the authors (available at https://predictdb.org/), derived from an elastic net regression model with transcriptome reference data from GTEx(v7). The most significant associations were observed for *DUS3L*, which showed significant associations (FDR < 0.05) with EWR.Total in four brain regions including amygdala (Z = -4.81, P = 4.18E-02), caudate basal (Z = -4.72, P = 4.18E-02), cerebellar hemisphere (Z = -4.69, P = 4.18E-02) and putamen (Z = -4.60, P = 4.77E-02) (see Table S2.1). The top 20 association results from S-PrediXcan are presented in Table 3 (see also Table S2 with top 100 associations).

**Table 3.**
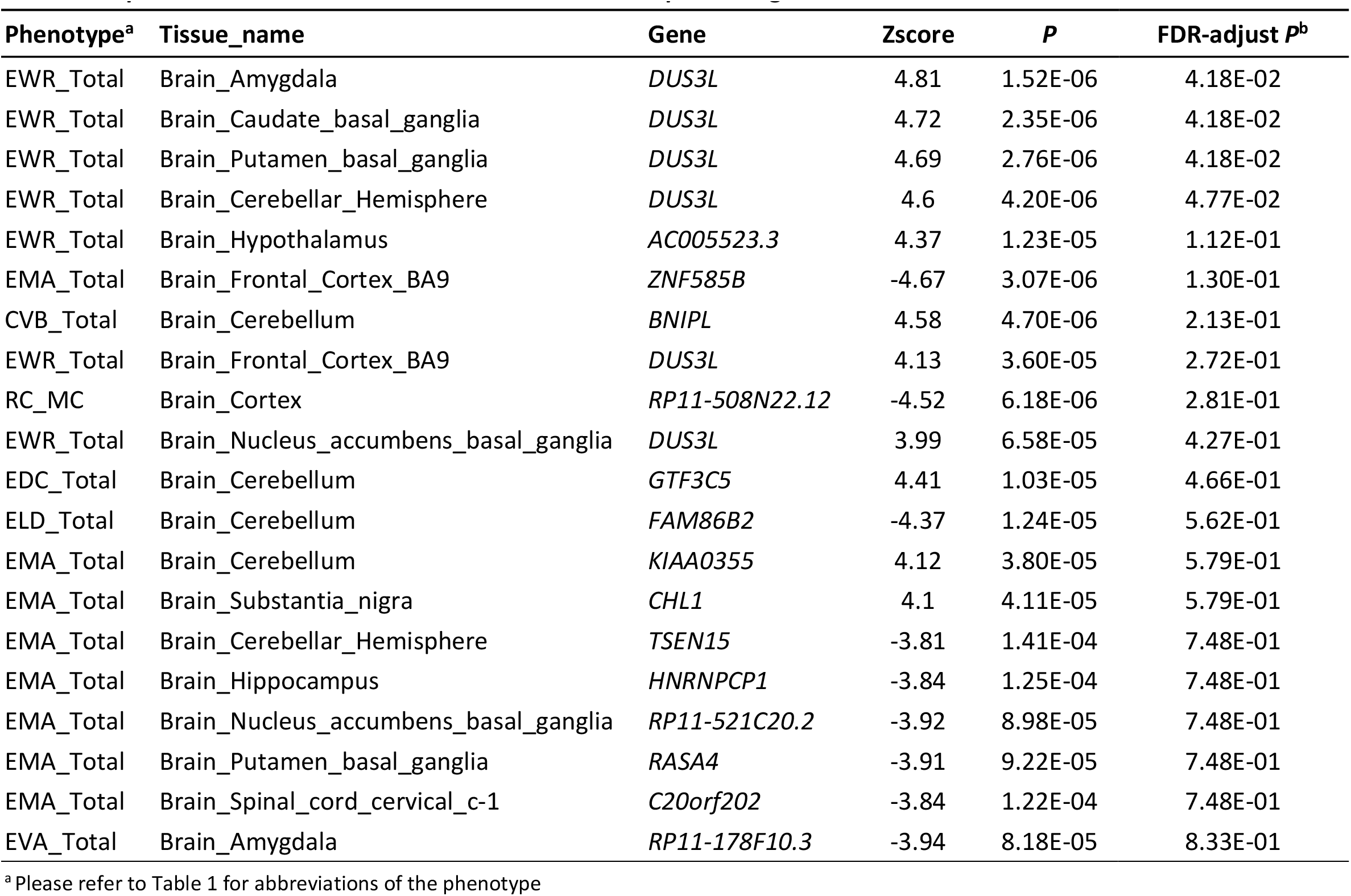
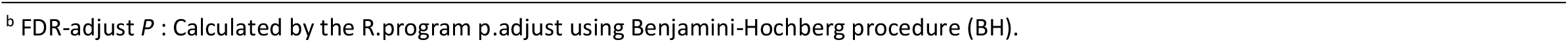
Top 20 S-Predixcan results after correction of multiple testing.

Furthermore, we also employed S-MulTiXcan to improve power by combining evidence of differential expression across all brain regions. We observed 185 significant gene-level associations (with FDR<0.05) by this approach and identified the best representative brain region (the region showing the strongest single-tissue association). The top 20 results are presented in Table 4 and fuller results in Table S3. We highlight a few findings here. The most significant S-Multixcan association was observed for gene *HSD3B7* with EVA_total (Hydroxy-Delta-5-Steroid Dehydrogenase, 3 Beta- And Steroid Delta-Isomerase 7; best brain region, Brain_Cortex; P = 1.12E-21). *HSD3B7* was also associated with other English literacy phenotypes, such as EVB, EVK, EVD, and EWR. For Chinese literacy skills, the most significant association was observed for gene *SEMA6C* (Semaphorin 6C; Brain_Cerebellar_Hemisphere; Z mean = - 1.81; P = 1.98E-16) with CVB_Total.

**Table 4.**
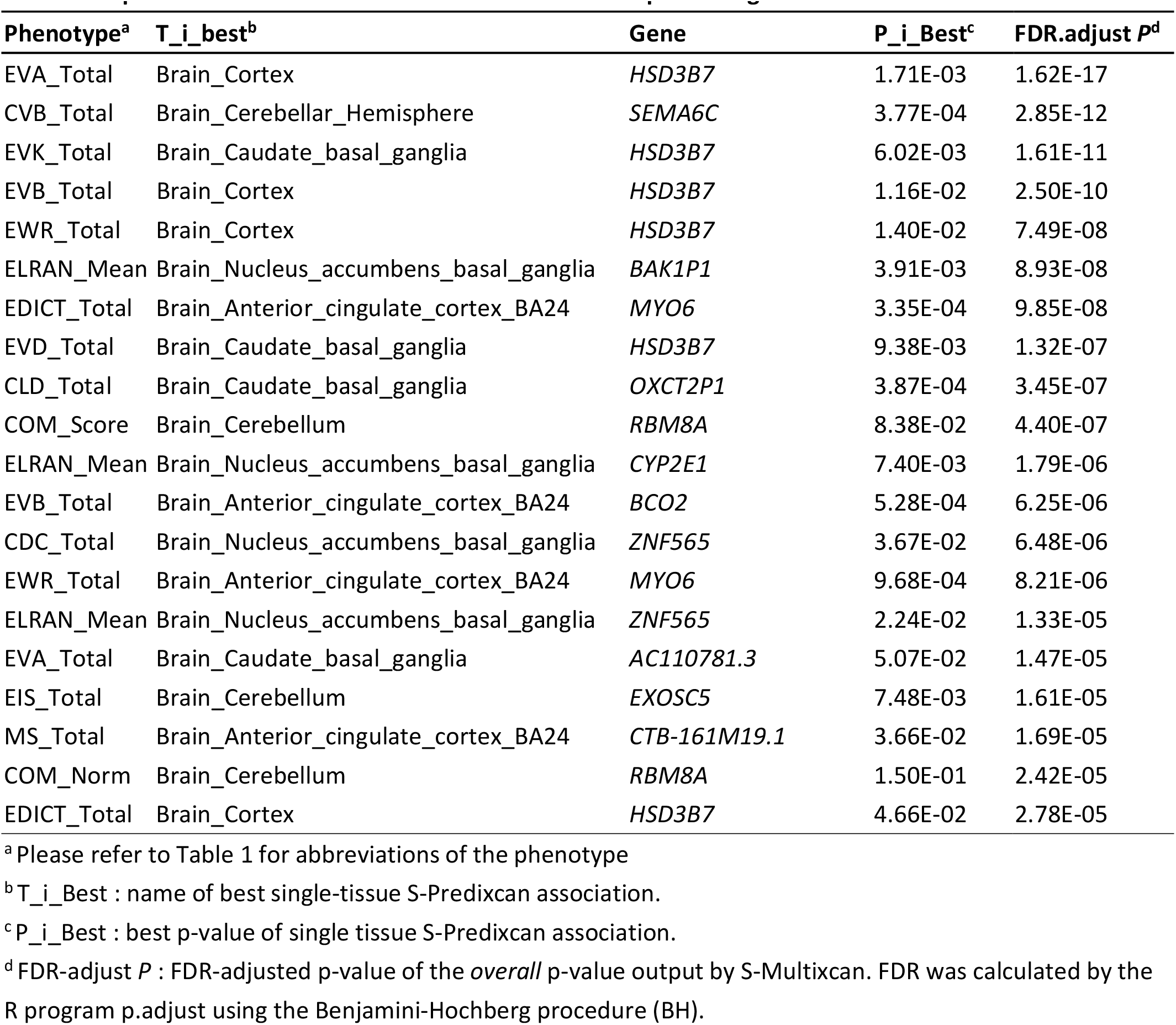
Top 20 S-Multixcan results after correction of multiple testing.

### Gene-based tests using MAGMA

We also conducted gene-based analyses using MAGMA, which aggregates SNP-level associations into a gene-level statistic. This approach considers the statistical significance of SNP-based test but does not explicitly consider how SNPs affect expression levels. The top 20 significant results from MAGMA are presented in Table 5 and full results are given in Table S4. We shall highlight several genes within the top-10 list here.

**Table 5.**
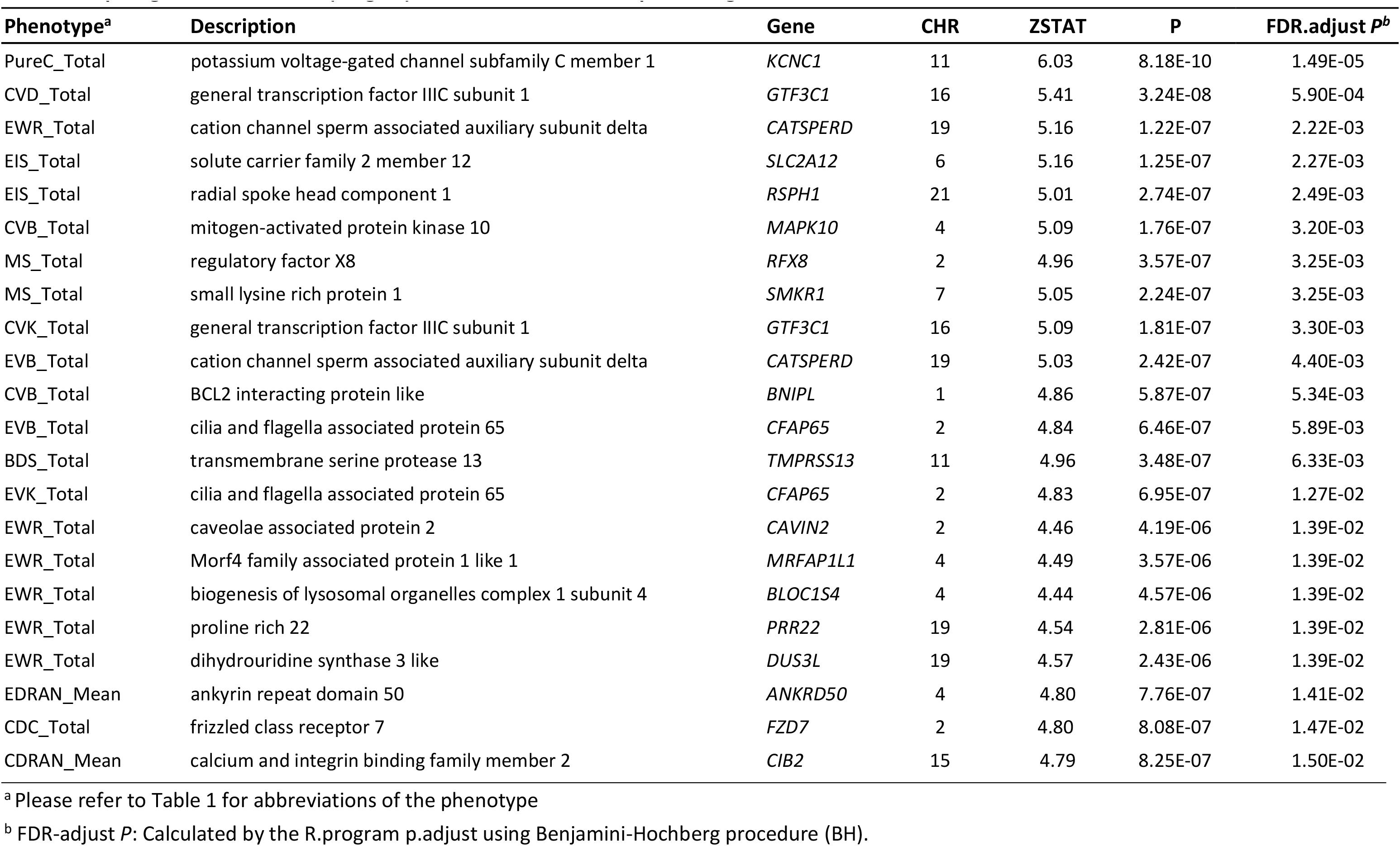
Top 20 gene-based results (Magma) after correction of multiple testing.

The most significant association was observed for *KCNC1* (potassium voltage-gated channel subfamily C member 1) with PureC_total (Z=6.03, FDR corrected p=1.49E-5). For English-related phenotypes, the most significant association was identified for gene *CATSPERD* (cation channel sperm associated auxiliary subunit delta) with EWR_Total (Z = 5.1632; FDR corrected *p* = 2.22E-03); the same gene was also associated with EVB_Total (Z = 5.0327; FDR corrected *p* = 4.40E-03). Two genes showed associations with EIS_Total, namely *SLC2A12* (solute carrier family 2 member 12; Z = 5.1581; FDR corrected *p* = 2.27E-03) and *RSPH1* (radial spoke head component 1; Z = 5.009; FDR-corrected *p* = 2.49E-03).

As for Chinese literacy skills, *GTF3C1* (general transcription factor IIIC subunit 1) was associated with CVD_Total (Z = 5.405; FDR corrected *p* = 5.90E-04) and CVK_Total (Z=5.09 ; FDR corrected *p* =3.03E-3); *MAPK10* (mitogen-activated protein kinase 10) was associated with CVB_Total (Z = 5.0938; FDR corrected *p* = 3.20E-03). As for morphosyntactic skills in Chinese, the genes *SMKR1*(small lysine rich protein 1; Z = 5.0475; FDR corrected *p* = 3.25E-03) and *RFX8* (regulatory factor X8; Z = 4.9575; FDR corrected *p* = 3.25E-03) were associated with MS_Total.

### Pathway enrichment analysis

To reveal relevant functional pathways, we conducted a self-contained gene-set analysis in GAUSS, testing 10679 canonical pathway and gene ontology (GO) gene sets from the MSigDB database. Full results with FDR<0.2 are shown in Tables S5.1 and S5.2. Table 6 and Table 7 summarize the top 20 pathway and GO analyses results. We also present the top two pathways and GO terms enrichment for every trait in Tables S5.3 and S5.4.

**Table 6.**
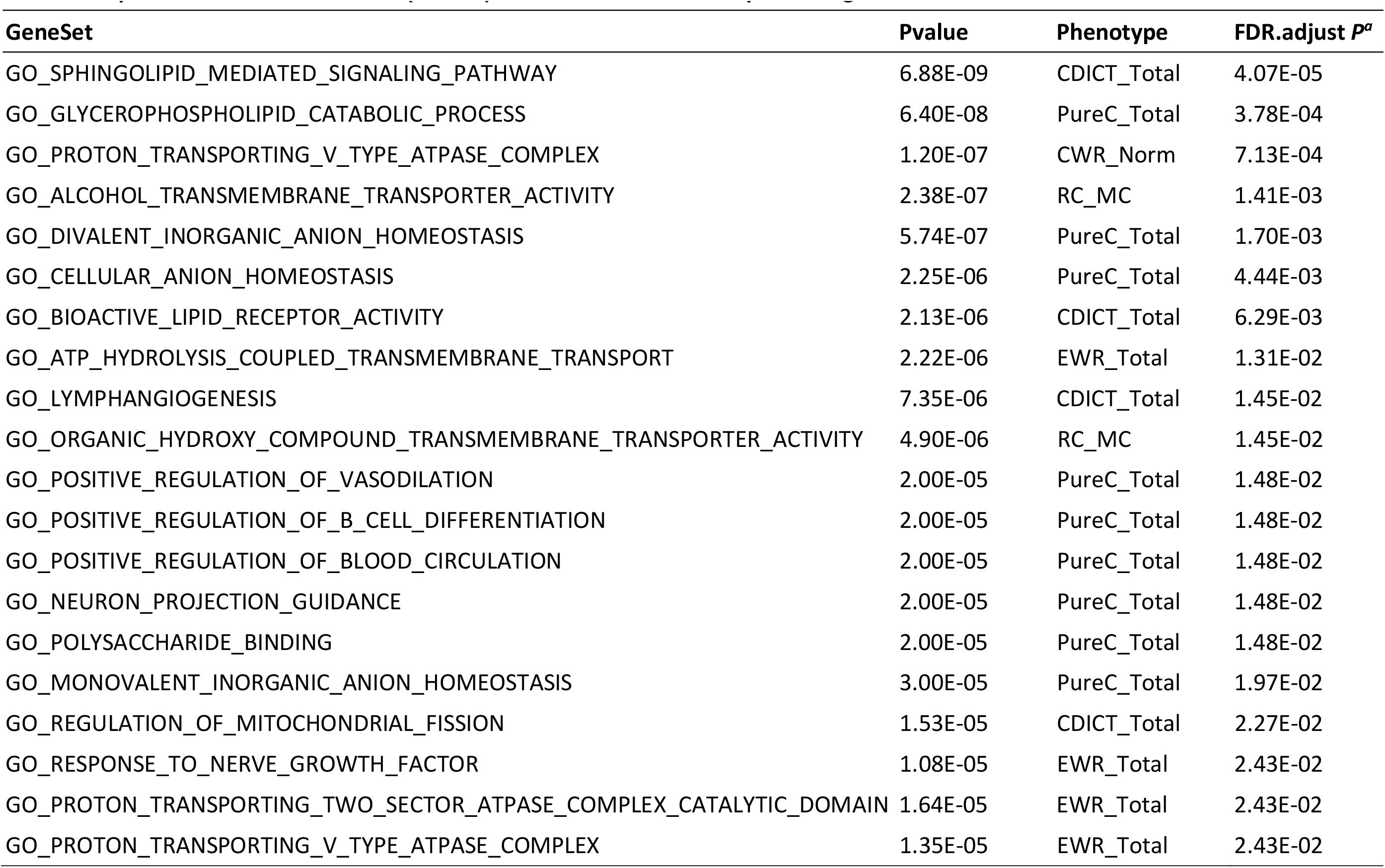
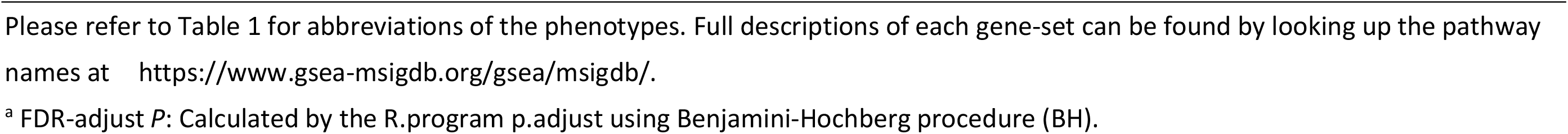
Top 20 GO enrichment results (GAUSS) after correction of multiple testing.

| GeneSet | Pvalue | Phenotype | FDR.adjust $P^a$ |
| --- | --- | --- | --- |
| GO_SPHINGOLIPID_MEDIATED_SIGNALING_PATHWAY | 6.88E-09 | CDICT_Total | 4.07E-05 |
| GO_GLYCEROPHOSPHOLIPID_CATABOLIC_PROCESS | 6.40E-08 | PureC_Total | 3.78E-04 |
| GO_PROTON_TRANSPORTING_V_TYPE_ATPASE_COMPLEX | 1.20E-07 | CWR_Norm | 7.13E-04 |
| GO_ALCOHOL_TRANSMEMBRANE_TRANSPORTER_ACTIVITY | 2.38E-07 | RC_MC | 1.41E-03 |
| GO_DIVALENT_INORGANIC_ANION_HOMEOSTASIS | 5.74E-07 | PureC_Total | 1.70E-03 |
| GO_CELLULAR_ANION_HOMEOSTASIS | 2.25E-06 | PureC_Total | 4.44E-03 |
| GO_BIOACTIVE_LIPID_RECEPTOR_ACTIVITY | 2.13E-06 | CDICT_Total | 6.29E-03 |
| GO_ATP_HYDROLYSIS_COUPLED_TRANSMEMBRANE_TRANSPORT | 2.22E-06 | EWR_Total | 1.31E-02 |
| GO_LYMPHANGIOGENESIS | 7.35E-06 | CDICT_Total | 1.45E-02 |
| GO_ORGANIC_HYDROXY_COMPOUND_TRANSMEMBRANE_TRANSPORTER_ACTIVITY | 4.90E-06 | RC_MC | 1.45E-02 |
| GO_POSITIVE_REGULATION_OF_VASODILATION | 2.00E-05 | PureC_Total | 1.48E-02 |
| GO_POSITIVE_REGULATION_OF_B_CELL_DIFFERENTIATION | 2.00E-05 | PureC_Total | 1.48E-02 |
| GO_POSITIVE_REGULATION_OF_BLOOD_CIRCULATION | 2.00E-05 | PureC_Total | 1.48E-02 |
| GO_NEURON_PROJECTION_GUIDANCE | 2.00E-05 | PureC_Total | 1.48E-02 |
| GO_POLYSACCHARIDE_BINDING | 2.00E-05 | PureC_Total | 1.48E-02 |
| GO_MONOVALENT_INORGANIC_ANION_HOMEOSTASIS | 3.00E-05 | PureC_Total | 1.97E-02 |
| GO_REGULATION_OF_MITOCHONDRIAL_FISSION | 1.53E-05 | CDICT_Total | 2.27E-02 |
| GO_RESPONSE_TO_NERVE_GROWTH_FACTOR | 1.08E-05 | EWR_Total | 2.43E-02 |
| GO_PROTON_TRANSPORTING_TWO_SECTOR_ATPASE_COMPLEX_CATALYTIC_DOMAIN | 1.64E-05 | EWR_Total | 2.43E-02 |
| GO_PROTON_TRANSPORTING_V_TYPE_ATPASE_COMPLEX | 1.35E-05 | EWR_Total | 2.43E-02 |

<sup>a</sup> FDR-adjust $P$ : Calculated by the R.program `p.adjust` using Benjamini-Hochberg procedure (BH).

**Table 7.** Top 20 Pathway enrichment results (GAUSS) after correction of multiple testing.

| **GeneSet** | **Pvalue** | **Phenotype** | **FDR adjust *P*<sup>a</sup>** |
| --- | --- | --- | --- |
| REACTOME_RNA_POL_III_TRANSCRIPTION | 3.36E-08 | WO_Total | 1.60E-04 |
| BIOCARTA_P35ALZHEIMERS_PATHWAY | 3.41E-07 | EWR_Total | 1.62E-03 |
| REACTOME_P2Y_RECEPTORS | 3.94E-07 | CVK_Total | 1.88E-03 |
| REACTOME_KINESINS | 7.07E-07 | BDS_Total | 3.37E-03 |
| STOSSI_RESPONSE_TO ESTRADIOL | 3.04E-06 | RC_MC | 1.45E-02 |
| IGLESIAS_E2F_TARGETS_DN | 4.29E-06 | CWR_Norm | 2.04E-02 |
| REACTOME_P2Y_RECEPTORS | 5.25E-06 | CVD_Total | 2.50E-02 |
| PID_S1P_META_PATHWAY | 9.02E-06 | CDICT_Total | 3.88E-02 |
| GOLUB_ALL_VS_AML_DN | 1.63E-05 | CDICT_Total | 3.88E-02 |
| BIOCARTA_AKAPCENTROSOME_PATHWAY | 2.00E-05 | CCR_Total | 4.76E-02 |
| BANDRES_RESPONSE_TO_CARMUSTIN_MGMT_48HR_UP | 2.00E-05 | CCR_Total | 4.76E-02 |
| LIM_MAMMARY_LUMINAL_PROGENITOR_UP | 2.00E-05 | EWR_Total | 4.76E-02 |
| BIOCARTA_BLYMPHOCYTE_PATHWAY | 1.17E-05 | CDC_Total | 5.55E-02 |
| BANDRES_RESPONSE_TO_CARMUSTIN_WITHOUT_MGMT_48HR_UP | 4.00E-05 | CCR_Total | 6.35E-02 |
| KEGG_PRIMARY_BILE_ACID_BIOSYNTHESIS | 3.00E-05 | EVb_Total | 6.35E-02 |
| RODRIGUES_NTN1_AND_DCC_TARGETS | 4.00E-05 | EVb_Total | 6.35E-02 |
| TERAMOTO_OPN_TARGETS_CLUSTER_7 | 4.00E-05 | EVb_Total | 6.35E-02 |
| KEGG_PROGESTERONE_MEDIATED_OOCYTE_MATURATION | 3.00E-05 | CVA_Total | 7.14E-02 |
| DUTERTRE ESTRADIOL_RESPONSE_6HR_UP | 2.00E-05 | CVA_Total | 7.14E-02 |
| SA_FAS_SIGNALING | 9.00E-05 | CDRAN_Mean | 7.14E-02 |

<sup>a</sup> FDR-adjust *P*: Calculated by the R.program p.adjust using Benjamini-Hochberg procedure (BH).

In pathway-based enrichment analysis of Chinese comprehension skills, the strongest association was observed for WO_Total with the Reactome RNA polymerase III transcription pathway (FDR corrected *p* = 1.60E-04). The second most significant association was observed for EWR_Total with the ‘Deregulation of CDK5 in Alzheimers Disease’ pathway (BioCarta) (FDR corrected *p* = 1.62E-03). Other pathways with the top five included the P2Y receptors (associated with CVK_total) and kinesins pathways (associated with BDS_total). GAUSS has also identified a collection of corresponding core genes (CS) for each pathway (please refer to Table S5).

In gene ontology (GO) enrichment analysis, the most significant enrichment was observed between CDICT_Total and sphingolipid-medicates signaling pathway (FDR corrected *p* = 4.07E-05). Other GO gene-sets within the top 5 (with respect to lowest p-values) included glycerophospholipid catabolic process, proton-transporting V-type ATPase complex, alcohol transmembrane transporter activity and divalent inorganic anion homeostasis. They were associated with PureC_total, CWR_norm, RC_MC and PureC_total, respectively. With regards to English literacy skills, we found that the GO gene-set ‘ATP hydrolysis coupled cation transmembrane transport’ (FDR corrected *p* = 1.31E-02) was the strongest association (with EWR_total). GAUSS selected 14 core genes for the gene set, in which one of them, *BLOC1S4*, was individually and significantly associated with EWR_Total (see TableS5.2).

### Look-up of loci reported in two previous GWAS of dyslexia and literacy traits

Among the 42 (*p* < 5e-08) and 161 loci (*p* < 1e-06) previously reported for dyslexia ^9^ and literacy phenotypes ^8^ respectively, none showed genome-wide significance in our analyses. To evaluate the *aggregate* effects of previously reported genes, we also performed a gene-set analysis, in which we considered the genome-wide significant genes from the two previous GWAS ^8 9^ as candidate ‘gene-sets’. An association was found between the literacy skills gene-set (derived from ref ^8^) with CVD_Total and CVK_Total (FDR < 0.05). Full results are reported in Table S8.

### Genetic overlap with neuropsychiatric phenotypes, cognitive performance (CP), and education attainment (EA)

Here we briefly describe several significant or suggestive findings (with FDR-corrected p<= 0.1) in PRS analysis. The ADHD polygenic score was associated with EVD_Total (FDR-corrected p= 0.026) at p-value threshold (pthres) *= 1e-06*. We also observed significant associations with PRS of EA. The strongest association was observed at pthres = 1e-02 with EIS_total (FDR-corrected p = 0.0035); EA PRS was also associated with EDICT, EMA, EVA, EVB, EVK and EWR (FDR corrected p<0.1) at one or more pthres levels. All the associations were in the positive direction. The CP PRS was positively associated with CVD, EDC, EVD, EVK, EWR and MS with FDR-corrected p<0.1, at one or more pthres levels.

For other neuropsychiatric phenotypes, SCZ PRS was associated with BDS_Total across multiple p-value thresholds. Also, we observed an association of MDD PRS with CVD and CVK, and an association of BP PRS with DS at FDR<=0.1. We did not observe significant associations otherwise and no associations were observed with ASD PRS.

We present in Figure 1 the results of a polygenic score analysis on different traits at the best pthres cutoff. Full PRS results across all pthres are reported in Table S6.

**Figure 1.**
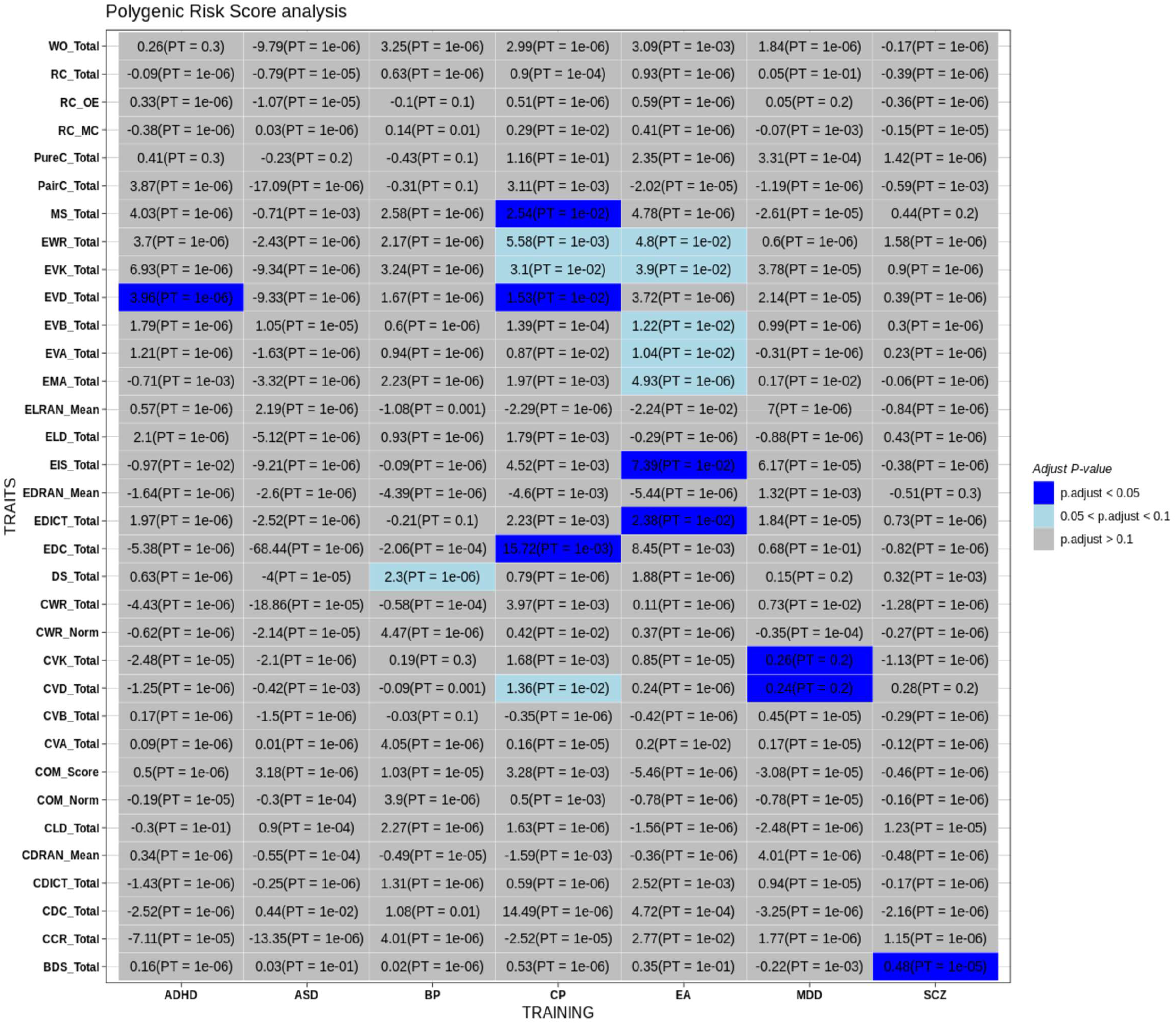
Results of polygenic risk score (PRS) analysis on the 34 language-related phenotypes analyzed in this study, with PRS constructed from external GWAS data of different neuropsychiatric disorders/traits (training set). The following neuropsychiatric disorders/traits were included: attention deficit hyperactivity disorder (ADHD), autism spectrum disorders (ASD), Education attainment (EA), cognitive performance (CP), schizophrenia (SCZ), bipolar disorder (BP) and major depressive disorder (MDD). In the heatmap, for each PRS analysis, we select the result with the lowest FDR-adjusted p-value (p.adjust), and show the regression coefficient in the graph. PT: the optimal p-value threshold at which the most significant association was observed

## Discussion

### Overview

In this study, we attempted to uncover the genetic basis of a comprehensive range of cognitive, literacy-related, and language-related phenotypes in Chinese and English. To gain robust insights into the genetic architecture of the above phenotypes, we carried out a GWAS within a group of Hong Kong children, mainly developing twins with Cantonese as their native language and English as their second language. To the best of our knowledge, this is the first GWAS to explore the genetic basis of a comprehensive set of literacy- and language-related traits in both Chinese and English in a Chinese population. Compared to the previous GWAS on language traits (see introduction), this study also covers the widest range of phenotypes, enabling a finer resolution into the genetic architecture of language abilities.

One distinct feature of this study is that we selected the subjects drawn from a large longitudinal project in Hong Kong, a city with a unique linguistic background due to its geographical location and political history ^45^. As such, our study represents the first attempt to assess the genetics of language and literacy skills of bilingual (Chinese and English) children systematically.

### Genes associated with literacy- or language-related phenotypes

For English literacy skills, the most significant association was observed for a SNP close to *MANEA* and *MANEA-AS1* (rs6905617) with English lexical decision. Interestingly, by a search of the GWAS catalog, we found that a variant in *MANEA* showed tentative association with general cognitive ability in a previous GWAS (*p*=5e-6) ^46^ ; genetic variants in *MANEA-AS1* may also be associated with behavioral inhibition ^47^. Regarding Chinese literacy skills, the SNP rs16848469 located in *TNR* (tenascin-R) was associated with CVD_Total and CVK_Total. *TNR* is primarily expressed in the central nervous system and plays a key role in human brain development by its involvement in axon growth and path finding^48^. Interestingly, variants in *TNR* have been reported to be associated with cognitive performance ^49 46^. In addition, a recent study ^50^ revealed that *TNR* showed a significant interaction (at *p*<5e-8) with total testosterone in affecting fluid intelligence in healthy adults. In addition, a recent GWAS on ADHD identified a variant in *TNR* as the top association achieving genome-wide significance ^51^. There were also case reports that deletions in *TNR* may be associated with intellectual disability and neurodevelopmental disorder ^52 53^. Moreover, animal studies showed that TNR-deficient mice had severe memory and coordination deficits ^54^.

In addition, we also observed that the SNP rs182977703 was associated with COM_Norm and CDICT_Total, which lies within the gene *SHTN1* (Shootin 1). The gene is involved in the generation of internal asymmetric signals necessary for neuronal polarization ^55^. Increased expression of *SHTN1* in hippocampal neurons may result in its accumulation in multiple neurites and formation of surplus axons^56^. Clinically, neurodevelopmental disorders such as intellectual disability (ID) may result from defects in neuronal polarity and migration ^57 58^. For example, the association of Shootin 1 with ID was supported by a whole-genome transcriptome analysis on ID patients ^59^. Another gene of interest in *PLXNC1*; variants in this gene have been reported to be associated (at *p*<1e-5) with multiple neuropsychiatric phenotypes such as major depression ^60^, Lewy body dementia ^61^, brain shape (segment 15 and 79) ^62^ and neuroticism^63^.

Several gene-based tests reached a significance level after FDR correction for reading and spelling measures. The most significant gene from MAGMA was *KCNC1*, which encodes a subunit of the KV3 voltage-gated K^+^ channels. Mutations in this gene were associated with a range of neurological disorders including epilepsy and also intellectual disability and cognitive decline in some patients ^64 65 66^. In terms of Chinese literacy skills, the most significant association signal was observed for gene *GTF3C1* (General Transcription Factor lllC Subunit 1) with CVD_Total. *GTF3C1* has been widely investigated on its interactive connections to other genes; for example, it is involved in networks pathologically related to neurodegenerative and Alzheimer’s disease ^67 68 69^. It has been shown that *GTF3C1* is involved in regulation of rearrangement of neuronal nuclear architecture following neuronal excitation ^70^. Of note, the nuclear architecture plays an important role in neural development and function ^71^. *CHL1* was another gene implicated from S-PrediXcan analysis, and variants in this gene have been reported to be associated with education attainment ^49^ and also mathematics abilities ^49^.

In addition, our results showed that *SLC2A12* appears to be associated with English comprehension skills. *SLC2A12* encodes GLUT12, a glucose transporter. It has been reported that amyloid-beta increases GLUT12 protein expression in the brain in mouse models, implicating an important role of this transporter in Alzheimer disease ^72^ and cognitive functioning.

### Polygenic score analysis

We discovered that some language traits were associated with PRS of psychiatric disorders, cognitive performance and educational attainment. We found that, for example, PRS for educational attainment and cognitive performance (derived from external GWAS data) were associated with various literacy traits. Our results were consistent with previous studies that have demonstrated shared genetic factors among childhood intelligence, educational attainment, and literacy skills.

For example, Luciano et al. (2017) ^73^ showed that PRS of word reading, general reading and spelling, as well as non-word repetition, were positively associated with educational attainment (college/university degree versus none), income and verbal-numerical cognitive test results. Moreover, in a GWAS by Price et al.,^74^ substantial genetic overlap was found between word reading and number of years of education (*R*^2^ = 0.07, *P* = 4.91 × 10^−48^) and intelligence score (*R*^2^ = 0.18, *P* = 7.25 × 10^−181^) in a population-based sample. In another recent study by Gialluisi et al. ^14^, risk of developmental dyslexia was also significantly associated with PRS of EA and intelligence. Combined with our current findings, these results provide evidence to support a partially shared genetic etiology among literacy skills, cognitive measures, and educational outcomes.

### Strengths and limitations

There are several strengths of our study. First of all, to the best of our knowledge, this is the first GWAS to investigate the genetic basis of a wide range of both Chinese and English literacy- and language-related skills in a Chinese population. Importantly, as reading and language comprehension are highly complex traits, here we performed detailed phenotyping to decipher the genetic basis of various different domains of these skills. On the other hand, previous studies largely followed another research strategy by focusing on a limited range of language phenotypes or binary outcomes. While it is also possible to only focus on a few selected phenotypes, for example, those with higher heritability (or by other criteria), such choice of phenotype may inevitably be somewhat arbitrary, and one may still discover variants of biological relevance for a trait with lower heritability. In addition, the SNP-based heritability, or the extent to which common variants contribute to a trait, is unknown for most phenotypes studied here. To enable a more comprehensive and unbiased examination of the genetic architecture of language/literacy-related traits, we have included a wide range of phenotypes in the current study. We also employed the FDR approach to account for multiple testing.

To gain deeper insights into the biological basis of the studied traits, we not only performed standard SNP-based tests but also gene-based (MAGMA, S-PrediXcan, S-MulTiXcan) and pathway-based analysis (GAUSS). This ‘multi-level’ approach helps to bridge the gap between SNP associations and biological mechanisms, thus enhancing our knowledge and understanding of reading and language. In addition to studying the associations between phenotypes and genetic factors, we performed PRS analysis to study the overlap of included phenotypes with other neuropsychiatric traits, which could provide insight into the genetic architecture of language-related traits.

Our study also has a few limitations. Our study is based on a Hong Kong Chinese sample (under a bilingual environment). It remains uncertain whether the genetic findings from the current study can be generalized to other populations. Further studies in other populations with different genetic and language backgrounds may be warranted. In a similar vein, the GWAS summary statistics of CP, EA and other psychiatric disorders were primarily derived from Europeans (due to lack of relevant data from Chinese populations), which may also attenuate the genetic overlap with the studied phenotypes in a Chinese population. Nevertheless, some studies (on other complex traits) have shown that genetic variants and PRS from Europeans may still be transferrable to Chinese ^75 76^. Also, here we employed the 1000-Genomes as reference for imputation, following the findings from Lin et al. ^26^ that satisfactory imputation performance in Chinese was achieved using the 1000G panel. In Lin et al.’s report, the mean imputation r^2^ in two Chinese cohorts were at or above ∼0.7 with MAF>1%, and were even better for higher MAF. At the time of this analysis, most established imputation servers (e.g. Michigan Imputation Server) does not contain Chinese-specific reference panels. Note that we also reported the imputation quality score (r^2^) for all reported variants for easy reference, and have removed variants with low imputation quality (r^2^<0.3).

In this study, we performed extensive and deep phenotyping covering most domains of Chinese and English literacy- and language-related skills. This GWAS covers the widest range of language phenotypes to date. However, admittedly, our sample size is relatively moderate and statistical power may be insufficient to detect variants of small effects. In addition, given that we only performed genetic analysis in a single sample and some phenotypes were studied for the first time (e.g. most phenotypes on Chinese reading/comprehension), we emphasize that further replications in other samples are required. As for the genetic analyses, this study focused on the contribution of common variants; rare variant association was not our focus and may require further sequencing studies. In addition, while we have performed further gene-based and pathway-based bioinformatics analyses, the findings are based on statistical associations and will require further experimental validations.

In summary, we have conducted the first GWAS on a comprehensive range of phenotypes on both Chinese and English abilities in a HK Chinese population. We discovered a number of novel genetic loci that may underlie these traits, and revealed genes and pathways that may be associated. We believe our work will be an important starting point and reference for further studies into the biological and genetic basis of language abilities, and ultimately such knowledge will be useful for the development of better treatment for children with specific reading disabilities.

## Data Availability

All data produced in the present work are contained in the manuscript.

https://drive.google.com/drive/folders/1AFZZrGI5zmijUo8M6KEtENC_P-ziMzIp?usp=sharing

## Acknowledgements

This study was partially supported by a Collaborative Research Fund (CRF) (C4054-17W) from the Research Grants Council. HCS was also supported by the Lo Kwee Seong Biomedical Research Fund.

## Supplementary Information

Supplementary information is available at the journal’s website and at https://drive.google.com/drive/folders/1AFZZrGI5zmijUo8M6KEtENC_P-ziMzIp?usp=sharing

